# Community responses during the early phase of the COVID-19 epidemic in Hong Kong: risk perception, information exposure and preventive measures

**DOI:** 10.1101/2020.02.26.20028217

**Authors:** Kin On Kwok, Kin-Kit Li, Henry Ho Hin Chan, Yuan Yuan Yi, Arthur Tang, Wan In Wei, Samuel Yeung Shan Wong

## Abstract

**Background:** Community responses are important for outbreak management during the early phase when non-pharmaceutical interventions are the major preventive options. Therefore, this study aims to examine the psychological and behavioral responses of the community during the early phase of the COVID-19 epidemic in Hong Kong.

**Method:** A cross-sectional online survey was launched within 36 hours after confirmed COVID-19 cases were first reported. Councilors of all 452 district council constituency areas were approached for survey dissemination. Respondent demographics, anxiety level, risk perception, sources to retrieve COVID-19 information, actual adoption and perceived efficacy of precautionary measures were collected.

**Result:** Analysis from 1715 complete responses indicated high perceived susceptibility (89%) and high perceived severity (97%). Most respondents were worried about COVID-19 (97%), and had their daily routines disrupted (slightly/greatly: 98%). The anxiety level, measured by the Hospital Anxiety and Depression Scale, was borderline abnormal (9.01). Nearly all respondents were alert to the disease progression (99.5%). The most trusted information sources were doctors (84%), followed by broadcast (57%) and newspaper (54%), but they were not common information sources (doctor: 5%; broadcast: 34%; newspaper: 40%). Only 16% respondents found official websites reliable. Enhanced personal hygiene practices and travel avoidance to China were frequently adopted (>77%) and considered effective (>90%). The adoption of social-distancing measures was lower (39%-88%), and their drivers for greater adoption include: being female (adjusted odds ratio [aOR]:1.27), living in the New Territories (aOR:1.32-1.55), perceived as having good understanding of COVID-19 (aOR:1.84) and being more anxious (aOR:1.07).

**Discussion:** Risk perception towards COVID-19 in the community was high. Most respondents are alert to the disease progression, and adopt self-protective measures. This study contributes by examining the psycho-behavioral responses of hosts, in addition to the largely studied mechanistic aspects, during the early phase of the current COVID-19 epidemic. The timely psychological and behavioral assessment of the community is useful to inform subsequent interventions and risk communication strategies as the epidemic progresses.

## INTRODUCTION

On 12 January 2020, the World Health Organization (WHO) declared the novel coronavirus which caused unknown pneumonia cases in Wuhan, Hubei Province, China since December 2019 as “2019-nCoV”, which was renamed by the International Committee on Taxonomy of Viruses as “SARS-CoV-2” on 11 February 2020. In parallel, the WHO formally named the disease caused by SARS-CoV-2 as “COVID-19”, short for Coronavirus Disease 2019. Back in late December 2019, a cluster of 27 pneumonia cases associated with SARS-CoV-2 with a common link to the Huanan Seafood Wholesale Market were reported [1], and the first death case attributable to SARS-CoV-2 occurred on 9 January 2020. Soon after the first global incidence was confirmed in Thailand on 12 January 2020, new cases were reported in different countries and were mostly associated with Wuhan travel history or residency. As of 20 February 2020, there have been 75465 confirmed cases in China, including 11633 severe cases and 2236 deaths [2]. In Hong Kong, the number of confirmed cases has risen to 68 on 20 February 2020 since the first local detection on 23 January 2020 [3].

Health officials have enacted interventions to slow transmission. In Hong Kong, adopted strategies include: border screening (measuring body temperature, imposing a health declaration form system, imposing a 14-day mandatory quarantine period on all individuals entering Hong Kong from the Mainland), social-distancing (border shutdown, reducing cross-border commuting services, deferring class resumption for schools governed by the Education Bureau, home-office arrangement for civil servants, suspension of public services from the Leisure and Cultural Services Department) and extending the Enhanced Laboratory Surveillance Program to adult patients with fever and mild respiratory symptoms presenting at accident and emergency departments or general out-patient clinics under the public sector.

To control this COVID-19 epidemic, much effort has been paid to identifying the etiological agent, epidemiological parameters such as incubation period [4], disease transmissibility [4, 5], clinical characteristics [6, 7], treatment options [8, 9], and route of transmission [10]. Although these information help devise optimal infection control strategies, such as contact tracing and follow-up isolation [11], they center purely on the mechanistic aspect of the disease.

The host’s behaviors are important for outbreak management, particularly during the early phase when no treatment or vaccination is available and non-pharmaceutical interventions (NPIs) are the only options. The efficacy of NPIs depends on the host’s degree of engagement and compliance in precautionary behaviors, such as wearing masks, hand hygiene and self-isolation. Whether individuals voluntarily engage in precautionary behaviors depends on their risk perception towards the current health threat. In fact, risk perception is a main theme in common health behavior theories, including Health Belief Model and Protection Motivation Theory. In addition, with advanced information technology in recent years comes the uncertainty of how risk perception is shaped by various information sources. Hong Kong’s past experience with outbreaks of novel pathogens (2003 Severe Acute Respiratory Syndrome (SARS), and 2009 Pandemic Influenza) also provides a reference point to evaluate the risk perceptions of the current COVID-19 epidemic.

In light of the importance of host behavior in mitigating transmission and the vision to inform policy formation in a timely manner, this study aims to examine risk perceptions and behavioral responses of the general community during the early phase of the COVID-19 epidemic. Considering the rapid development of the epidemic during the survey period and the potential variability in the adoption of preventive measures among hosts, this study also examines the temporal changes in anxiety, the factors associated with adoption of preventive measures and their sources of information gathering.

## METHODS

### Subject recruitment

A cross-sectional online survey was conducted within 36 hours after the first confirmed COVID-19 case was reported in Hong Kong. To ensure good coverage of the general community in Hong Kong, chairpersons and vice-chairpersons of all eighteen district councils and all individual councilors of the 452 District Council Constituency Areas (DCCAs) were approached by electronic mails and their contact numbers listed in the District Council websites (https://www.districtcouncils.gov.hk/index.html) for survey dissemination. District councilors were invited to share our survey link and promotion messages on their webpages, social media platforms or any channels which they usually use to convey information to their targeted residents, but in general there was no restriction on their dissemination. Individuals who were aged 18 or above, understood Chinese and lived (on average) over five days per week in Hong Kong in the last month are eligible to participate. Respondents were compensated with a HKD10 cash coupon if they indicated willingness for receipt. To avoid duplicated responses from the same respondent, the survey could only be taken once from the same electronic device.

### Respondent characteristics

Respondents were asked about their demographics (including sex, age, living district, education attainment, household income), self-perceived health status, travel history in the past month, occurrence of respiratory symptoms in the past fourteen days and anxiety level using the Hospital, Anxiety and Depression scale - Anxiety (HASD-A) (0-7 = Normal; 8-10 = Borderline abnormal; 11-21 = Abnormal). Although HADS-A is intended for screening clinically significant anxiety symptoms in clinical populations, many studies have showed that it is valid for community populations [12, 13], including employees [14], general population aged 65-80 years in Sweden [15], an Italian community sample aged 18-85 years [16]. As a complementary measure, the state anxiety level of a subset of respondents was assessed with the validated State Trait Anxiety Inventory (STAI).

### Risk perception

Risk perception towards COVID-19 was measured by two psychological dimensions: (i) perceived susceptibility, and (ii) perceived severity. The first dimension was proxied by how likely one considered oneself (his/her families) would be infected with COVID-19 if no preventive measure was taken. The second dimension was proxied by how one rated the seriousness of symptoms caused by COVID-19, their perceived chance of having COVID-19 cured and that of survival if infected with COVID-19. Subjects were also asked to rate the relative severity of COVID-19 compared with common non-communicable diseases (NCDs) and previous outbreaks by novel pathogens in Hong Kong. Responses were captured with a five-point Likert scale.

### Information exposure

Respondents were asked about the sources from which they obtained information about COVID-19, and how much they trust those sources. They were also asked about the types of information that they wanted to receive.

### Preventive measures

Respondents were asked whether they performed precautionary measures and what their perceived efficacy of those measures are. Three types of precautionary measures were considered: hygienic practices, social distancing and travel avoidance.

### Ethics consideration

This study has been approved by the Survey and Behavioral Research Ethics Committee of The Chinese University of Hong Kong.

### Patient and Public Involvement Statement

It was not appropriate or possible to involve patients or the public in the design, or conduct, or reporting, or dissemination plans of this research.

### Statistical analysis

Frequency and proportions of responses were tabulated. Demographics of respondents were compared to the 2016 population by-census in Hong Kong with Cohen’s w effect size (small: 0.1; medium: 0.3; large: 0.5) [17]. Regression models were used to test for temporal change in anxiety level and to identify factors associated with greater adoption of social-distancing preventive measures. The latter is proxied by adopting five or more social-distancing precautionary measures. Variables that appears to be associated (p<0.2) in the univariate analysis are considered in the multivariate analysis. The final model is determined by stepwise selection. Adjusted odds ratio (aOR) and 95% confidence interval (CI) are estimated. Candidate variables include: demographics of respondents, self-perceived health status, travel history and anxiety level. A statistical significance of 0.05 was specified. Analysis was performed in R.

## RESULTS

The survey was conducted from 24 January 2020 to 13 February 2020 (Figure 1). Our survey period covers important clinical incidences, including first local death case and first overseas death case (Philippines), and social incidences, including healthcare workers on strike to call for entire border shutdown. It was also amid of the start-up of large-scale social-distancing interventions, including halt of sales of high-speed rail tickets to and from Wuhan, closure of public cultural and leisure facilities and deferral of school resumption. Meanwhile, alongside the launch of this survey was the escalating official threat tone on COVID-19: The WHO declared the COVID-19 epidemic as a public health emergency of international concern, with Hong Kong activated the emergency response level.

**Figure 1.**
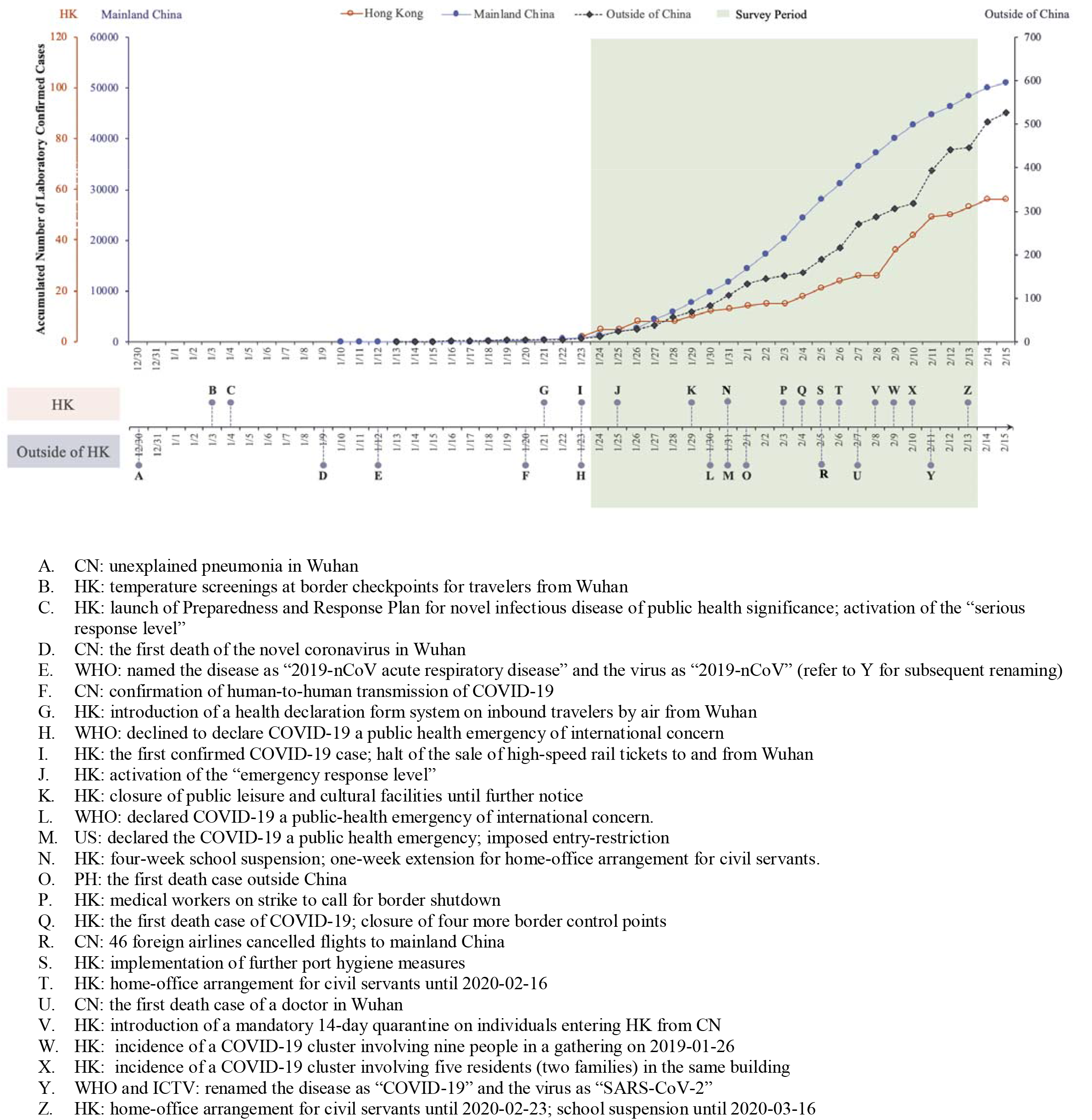
Laboratory confirmed cases and chronology of major events of COVID-19 Abbreviations: China (CN); Hong Kong (HK); International Committee on Taxonomy of Viruses (ICTV); the Philippines (PH); the United States (US); World Health Organization (WHO)

### Respondent characteristics

Complete data from 1715 respondents were analyzed. Table 1 shows the demographics of respondents. Many of the respondents are female (69%; 1176/1715), of young age (18-44 years) (80%; 1380/1715), working population (68%; 1168/1715). The study sample is comparable to the population in terms of residential district (effect size=0.27). Table 2 shows the background health conditions and travel history of respondents. The majority perceived their health status as good or very good (78%; 1331/1715), a quarter of them experienced respiratory symptoms in the past 14 days (423/1715) and travelled outside Hong Kong in the previous month (408/1715). Among the 408 respondents who were abroad, at least 24% of them (96-109) went to the Mainland China excluding Macau.

**Table 1.**
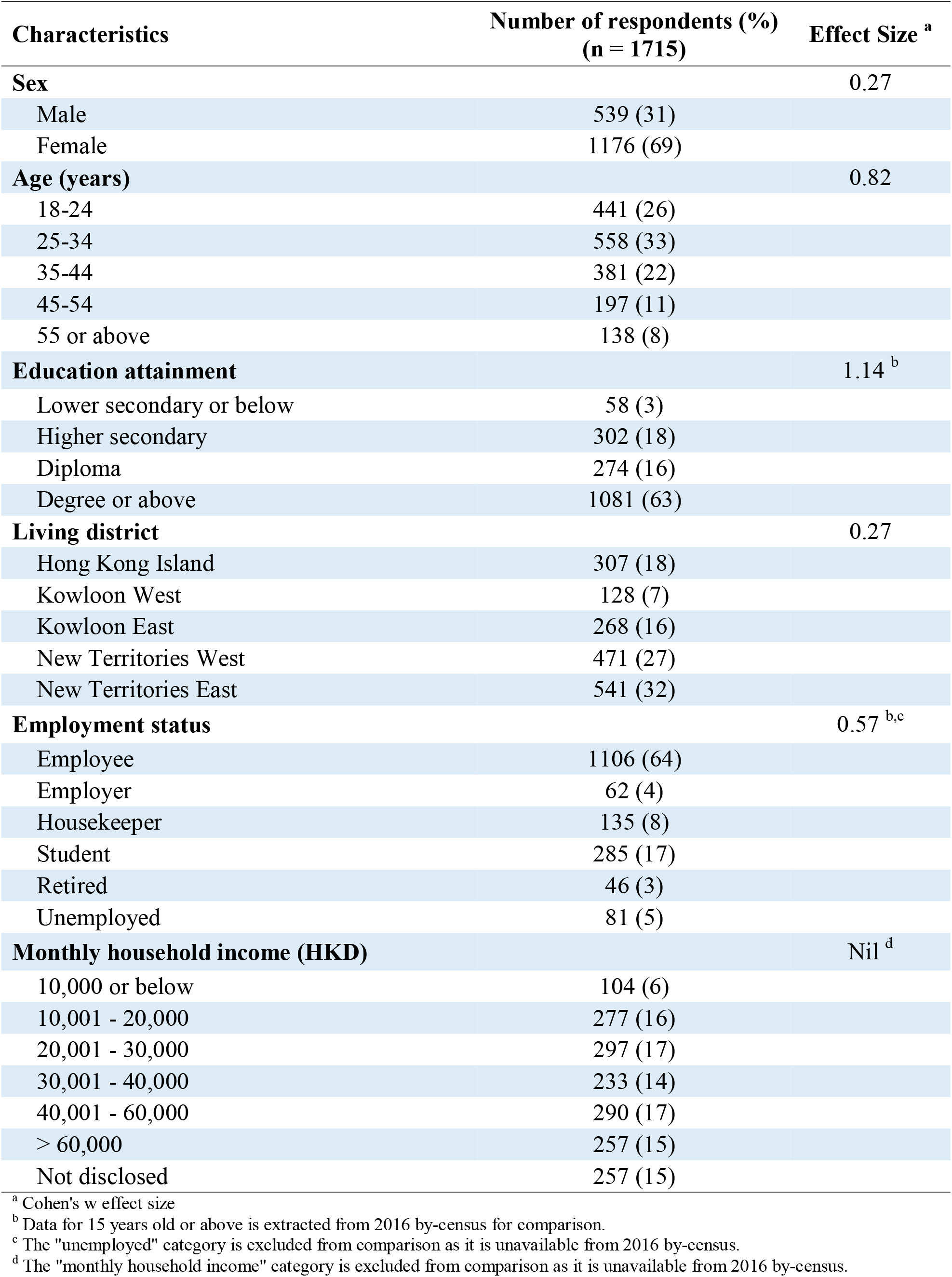
Respondent characteristics

| Characteristics | Number of respondents (%)<br>(n = 1715) | Effect Size <sup>a</sup> |
| --- | --- | --- |
| **Sex** |  | 0.27 |
| Male | 539 (31) |  |
| Female | 1176 (69) |  |
| **Age (years)** |  | 0.82 |
| 18-24 | 441 (26) |  |
| 25-34 | 558 (33) |  |
| 35-44 | 381 (22) |  |
| 45-54 | 197 (11) |  |
| 55 or above | 138 (8) |  |
| **Education attainment** |  | 1.14 <sup>b</sup> |
| Lower secondary or below | 58 (3) |  |
| Higher secondary | 302 (18) |  |
| Diploma | 274 (16) |  |
| Degree or above | 1081 (63) |  |
| **Living district** |  | 0.27 |
| Hong Kong Island | 307 (18) |  |
| Kowloon West | 128 (7) |  |
| Kowloon East | 268 (16) |  |
| New Territories West | 471 (27) |  |
| New Territories East | 541 (32) |  |
| **Employment status** |  | 0.57 <sup>b,c</sup> |
| Employee | 1106 (64) |  |
| Employer | 62 (4) |  |
| Housekeeper | 135 (8) |  |
| Student | 285 (17) |  |
| Retired | 46 (3) |  |
| Unemployed | 81 (5) |  |
| **Monthly household income (HKD)** |  | Nil <sup>d</sup> |
| 10,000 or below | 104 (6) |  |
| 10,001 - 20,000 | 277 (16) |  |
| 20,001 - 30,000 | 297 (17) |  |
| 30,001 - 40,000 | 233 (14) |  |
| 40,001 - 60,000 | 290 (17) |  |
| > 60,000 | 257 (15) |  |
| Not disclosed | 257 (15) |  |

<sup>a</sup> Cohen's w effect size
<sup>b</sup> Data for 15 years old or above is extracted from 2016 by-census for comparison.
<sup>c</sup> The "unemployed" category is excluded from comparison as it is unavailable from 2016 by-census.
<sup>d</sup> The "monthly household income" category is excluded from comparison as it is unavailable from 2016 by-census.

**Table 2.** Background health conditions and travel history of respondents

| Characteristics | Number of respondents (%)<br>(n = 1715) |
| --- | --- |
| **Self-perceived health status** |  |
| Very good / good | 1331 (78) |
| Fair | 352 (21) |
| Very bad / bad | 32 (2) |
| **Medical consultation<sup>a</sup> in the past 14 days** |  |
| Yes | 293 (17) |
| No | 1422 (83) |
| **Presence of respiratory symptoms in the past 14 days** |  |
| Yes | 423 (25) |
| No | 1292 (75) |
| **Leave Hong Kong in the previous month** |  |
| Yes<sup>b</sup> | 408 (24) |
| No | 1307 (76) |
| **Regular visitors to the Mainland China** |  |
| Yes<sup>c</sup> | 46 (3) |
| No | 1669 (97) |

<sup>a</sup> Both Chinese and Western medical consultations are included.
<sup>b</sup> Multiple destinations are allowed. Number of respondents (out of 408) who indicated travel outside Hong Kong in the previous month: outside China (294), China - Guangdong province (96), China - other province (13), Macau (29).
<sup>c</sup> Number of respondents (out of 46) who indicated regular visit to the Mainland China: daily (4), weekly (7), monthly (21), quarterly (4), and at most quarterly (10).

### Risk perception

Table 3 shows the perceived susceptibility and perceived severity towards COVID-19 among respondents. Most respondents regarded themselves as likely to be infected with COVID-19 (very likely/likely: 89%), and most considered the symptoms of COVID-19 (if infected) as serious (very serious/serious: 97%). Less than a quarter of the respondents thought that it was likely to have COVID-19 cured (if infected) (15%), and only 18% thought that it was likely to survive through COVID-19. When referencing to existing diseases (Table 4), almost all respondents (>98%) consider equivalent disease severity between COVID-19 and SARS. This magnitude was similar to other deadly NCDs (85%-94%), but much higher than the annual seasonal influenza (66%).

**Table 3.** Risk perception of the community towards COVID-19

|  | Number (%) |  |  |  |  |
| --- | --- | --- | --- | --- | --- |
|  | Level 1 | Level 2 | Level 3 | Level 4 | Level 5 |
| **Perceived susceptibility**<br>**(assuming no preventive measure)** |  |  |  |  |  |
| How likely you will be infected <sup>a</sup> | 776 (45) | 751 (44) | 160 (9) | 23 (1) | 5 (0) |
| How likely your families will be infected <sup>a</sup> | 924 (54) | 660 (38) | 113 (7) | 14 (1) | 4 (0) |
| **Perceived severity** |  |  |  |  |  |
| Seriousness of symptoms caused by SARS-CoV-19 <sup>b</sup> | 1102 (64) | 569 (33) | 33 (2) | 7 (0) | 4 (0) |
| Chance of having COVID-19 cured <sup>c</sup> | 190 (11) | 552 (32) | 708 (41) | 239 (14) | 26 (2) |
| Chance of survival if infected with COVID-19 <sup>c</sup> | 136 (8) | 476 (28) | 788 (46) | 290 (17) | 25 (1) |

<sup>a</sup> Level 1 = Very likely; Level 2 = Likely; Level 3 = Neutral; Level 4 = Unlikely; Level 5 = Very unlikely
<sup>b</sup> Level 1 = Very serious; Level 2 = Serious; Level 3 = Neutral; Level 4 = Not serious; Level 5 = Not serious at all
<sup>c</sup> Level 1 = Very low; Level 2 = Low; Level 3 = Neutral; Level 4 = High; Level 5 = Very high

**Table 4.** Comparison of disease severity

| **Diseases** | **Very bad** | **Bad** | **Neutral** | **Not bad** | **Not bad at all** |
| --- | --- | --- | --- | --- | --- |
| **Emerging infectious disease** |  |  |  |  |  |
| COVID-19 | 1545 (90) | 150 (9) | 15 (1) | 0 (0) | 1 (0) |
| **Existing infectious diseases** |  |  |  |  |  |
| SARS | 1551 (91) | 133 (8) | 21 (1) | 2 (0) | 4 (0) |
| 2009 pandemic influenza | 604 (35) | 889 (52) | 172 (10) | 40 (2) | 6 (0) |
| Seasonal influenza | 191 (11) | 948 (55) | 311 (18) | 251 (15) | 10 (1) |
| **Non-communicable diseases** |  |  |  |  |  |
| Diabetes | 659 (39) | 804 (47) | 188 (11) | 51 (3) | 9 (1) |
| Cancer | 1432 (84) | 215 (13) | 45 (3) | 11 (1) | 8 (0) |
| Heart disease | 1123 (66) | 502 (29) | 66 (4) | 17 (1) | 3 (0) |
| Acquired immune deficiency syndrome | 1354 (79) | 257 (15) | 69 (4) | 22 (1) | 9 (1) |

Most respondents were worried about COVID-19 (97%; 1667/1715), and they claimed that their daily routines were slightly (42%; 727/1715) or greatly (56%; 955/1715) disrupted. The average HADS-A score is 9.01 (95% CI: 8.44, 9.59); while the average score of state anxiety by STAI, from 804 complete responses, is 2.00 (95% CI: 1.46, 2.55). A significantly increasing time trend in HADS-A score is identified (p<0.05) (Figure 2).

**Figure 2.**
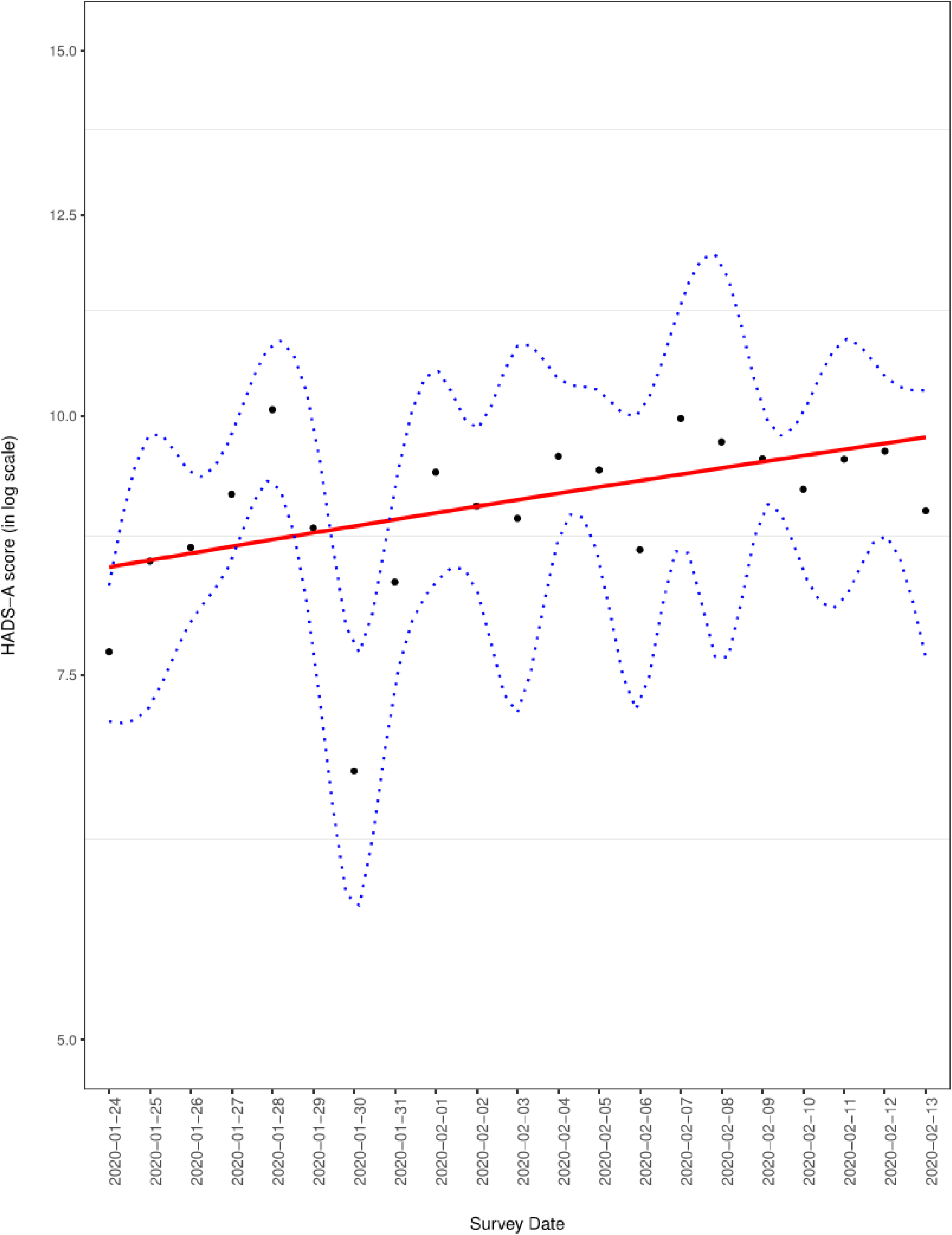
Time trend of HADS-A score

### Information exposure

Nearly all respondents were continuously alert to the disease progression of COVID-19 (99.5%; 1707/1715) and actively searched for related information (83%; 1431/1715). Table 5 lists the types of COVID-19 information wanted by the 1639 (96%) respondents who indicated such need. Information which respondents were most interested were: distribution of cases (92%), number of infected individuals (91%), infection control interventions undertaken by local officials (88%), and preventive measures (87%).

**Table 5.** Information wanted by the respondents

| **Information you want to receive about COVID-19** | **Number (%)**<br>**(n = 1639)** |
| --- | --- |
| Distribution of cases | 1506 (92) |
| Number of people infected | 1497 (91) |
| Interventions of Hong Kong government | 1450 (88) |
| Preventive measures | 1424 (87) |
| Disease progression | 1327 (81) |
| Symptoms/how to know if one is infected | 1310 (80) |
| Interventions of international organizations | 1182 (72) |
| What to do if infected | 1087 (66) |
| Impact on risk groups | 1073 (65) |
| Risks and consequences | 1061 (65) |
| Interventions of Chinese government | 1010 (62) |

Figure 3 shows the sources from which respondents obtained information about COVID-19, and how well the information sources were trusted. The most trusted sources were doctors (84%; very reliable/reliable: 1449/1715), but only 5% (87/1715) respondents could obtain information from them. The next two most trusted sources were broadcast (57%) and newspaper (54%), but they were used by less than 40% of the respondents. On the other hand, the two most common information sources were social platforms (94%; 1608/1715) and websites (regardless of official or unofficial) (90%; 1539/1715), but they were rated as reliable or very reliable by only 26% and 16%-23% of the respondents respectively. Only 16% (269/1715) of respondents found information from official websites reliable or very reliable.

**Figure 3.**
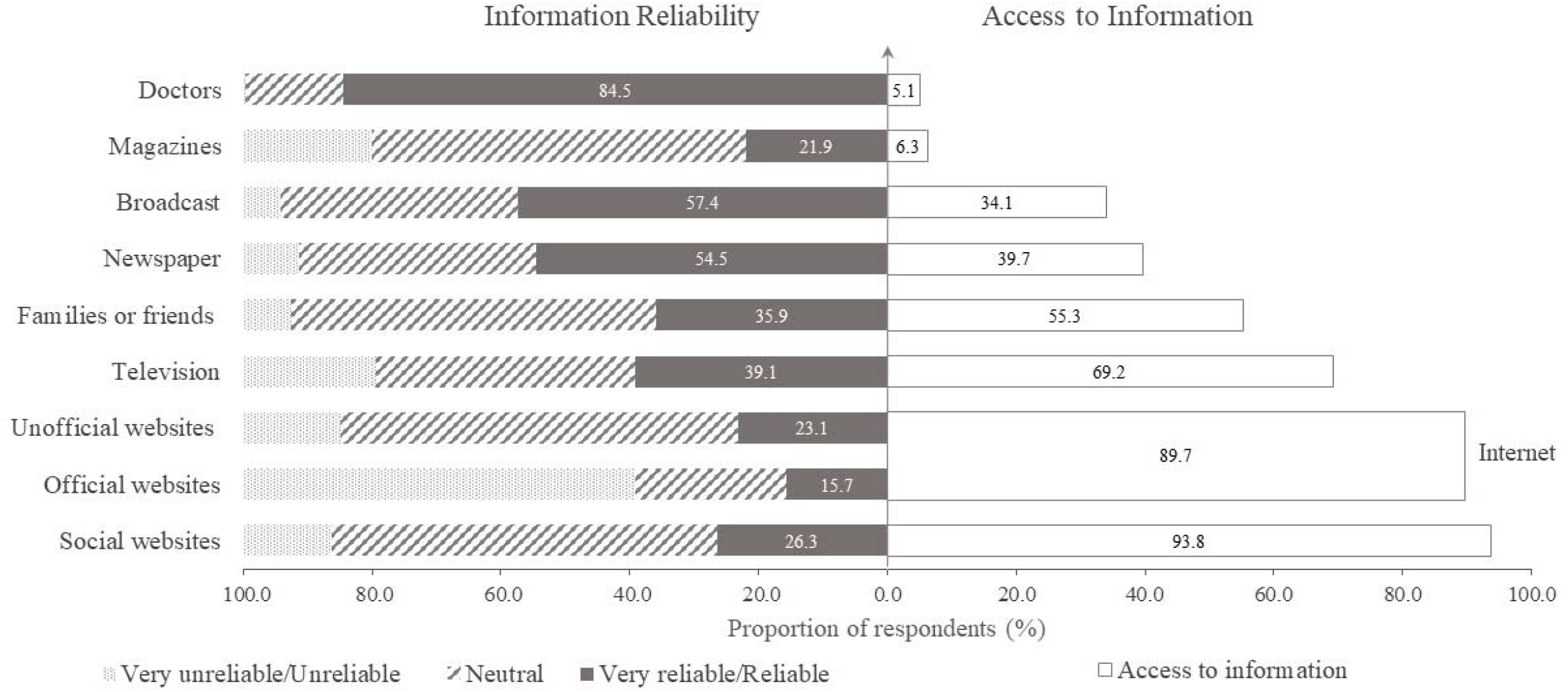
Information reliability and the access to information of COVID-19

### Preventive measures

Figure 4 shows the adoption of precautionary measures by respondents and their perceived efficacy. Enhanced personal hygiene practices (including wearing masks, cleaning hands and better coughing and sneezing etiquette) and avoid travelling to Mainland China were adopted by most respondents (>89%), and these practices were considered very effective or effective (>90%). For social-distancing measures, although they were considered useful in preventing COVID-19 (very effective/effective: 70%-93%), their actual adoption was lower (range: 39%-88%).

**Figure 4.**
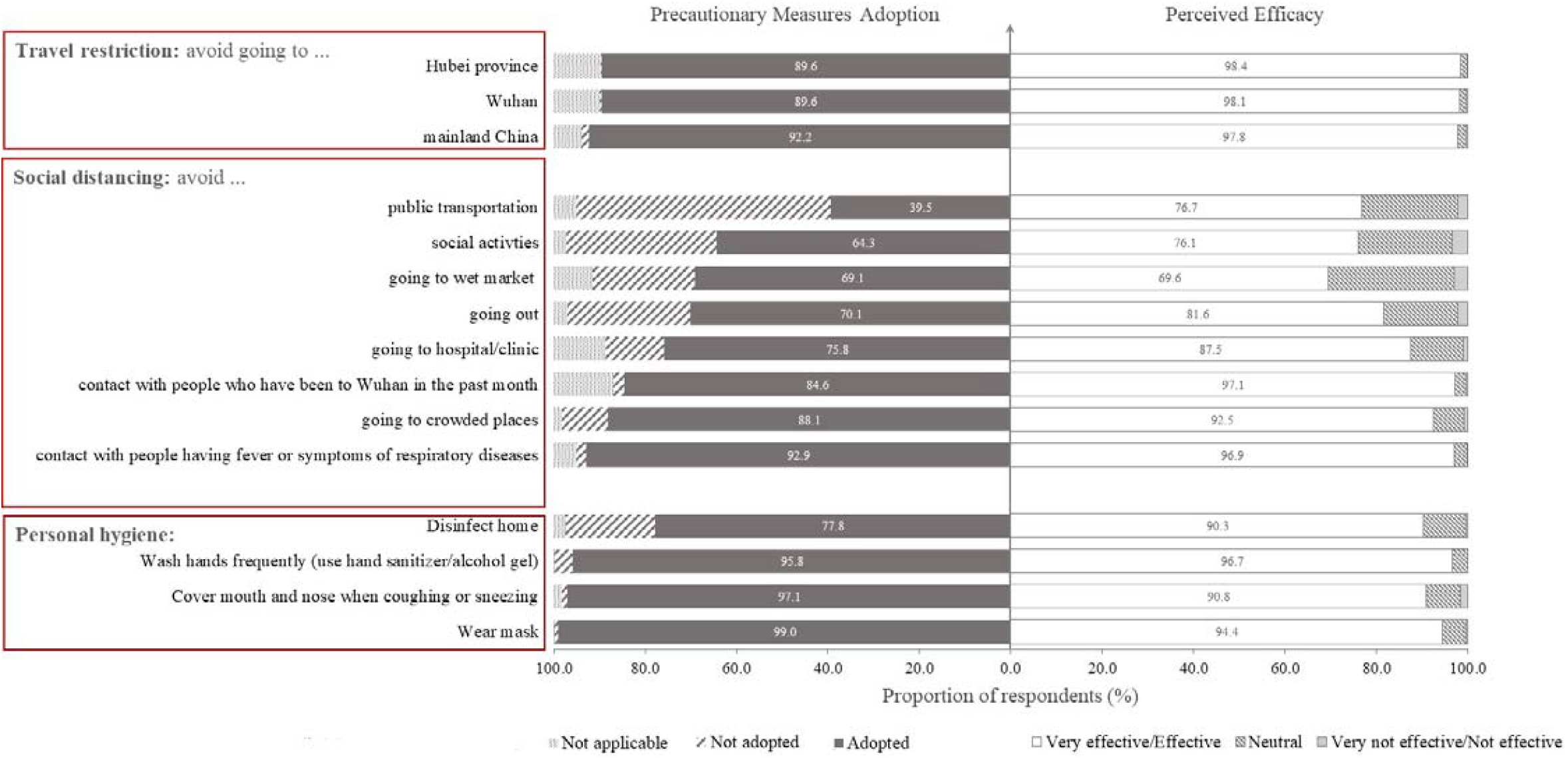
Perceived efficacy and actual adoption of precautionary measures to prevent transmission and contracting COVID-19

Table 6 shows the regression analysis results for greater adoption (five or more) of social-distancing interventions during the early phase of this COVID-19 epidemic. Being female (aOR:1.27; 95% CI:1.02,1.58), living in the NT (aOR:1.32-1.55), perceived as having good understanding of COVID-19 (aOR:1.84; 95% CI:1.29,2.63), being more anxious (aOR:1.07; 95% CI: 1.04,1.10) were positively associated with greater adoption.

**Table 6.** Factors associated with greater adoption of social-distancing interventions during the early phase of the COVID-19 epidemic in Hong Kong

| Characteristics | aOR (95% CI) | p-value |
| --- | --- | --- |
| **Sex** |  |  |
| Male | - |  |
| Female | 1.27 (1.02 , 1.58) | 0.03 |
| **Age (years)** |  |  |
| 18-24 | - |  |
| 25-34 | 1.21 (0.86 , 1.69) | 0.28 |
| 35-44 | 1.06 (0.74 , 1.53) | 0.75 |
| 45-54 | 1.08 (0.71 , 1.66) | 0.71 |
| 55 or above | 0.61 (0.36 , 1.02) | 0.06 |
| **Living district** |  |  |
| Hong Kong Island | - |  |
| Kowloon West | 0.97 (0.63 , 1.50) | 0.89 |
| Kowloon East | 0.93 (0.66 , 1.32) | 0.69 |
| NT West | 1.32 (0.98 , 1.79) | 0.07 |
| NT East | 1.55 (1.15 , 2.08) | <0.01 |
| **Presence of respiratory symptoms in the past 14 days** |  |  |
| No | - |  |
| Yes | 0.82 (0.65 , 1.04) | 0.1 |
| **Leave Hong Kong in the previous month** |  |  |
| No | - |  |
| Yes | 0.71 (0.56 , 0.90) | <0.01 |
| **Regular visit to mainland China** |  |  |
| No | - |  |
| Yes | 0.49 (0.25 , 0.97) | 0.04 |
| **Perceived understanding to COVID-19** |  |  |
| Not well / not well at all | - |  |
| Neutral | 1.10 (0.78 , 1.56) | 0.58 |
| Well / very well | 1.84 (1.29 , 2.63) | <0.01 |
| **Work status in the past seven days** |  |  |
| Employee | - |  |
| Employer | 1.77 (1.03 , 3.05) | 0.04 |
| Unemployed | 1.62 (1.01 , 2.60) | 0.05 |
| Housekeeper | 2.13 (1.42 , 3.21) | <0.01 |
| Student | 1.03 (0.71 , 1.50) | 0.86 |
| Retired | 2.08 (1.03 , 4.19) | 0.04 |
| **HADS-A score** | 1.07 (1.04 , 1.10) | <0.01 |

## DISCUSSION

This study provides timely assessment of the risk perception, information exposure and adoption of precautionary measures during the initial phase of the COVID-19 epidemic in Hong Kong. Despite disease uncertainty (including transmissibility, route of transmission and pathogenicity) at the early stage, individuals in the community had high perceived risk towards COVID-19 at large, viz: high perceived susceptibility and high perceived severity. A slightly increasing general anxiety level was observed over the three-week study period. Enhanced personal hygiene and travel avoidance were adopted by nearly all respondents, higher propensity of adopting greater degree of social-distancing measures were associated with being female, living in the NT, perceived as having good understanding of COVID-19 well, work status except students and being more anxious.

Our results have several immediate and significant public health implications. First, our results provide the baseline psychological and behavioral responses of the community against which current infection control strategies fit in. With the high perceived risk and large proportion of individuals adopting preventive measures in the community at the beginning, during which the accumulated number of local cases is 68 (as of 21 February 2020) with a significant initial portion of them being imported cases [18], we have an edge to block local transmission. This suggests that efforts to curb imported cases were efficient at the early phase of this outbreak. Following the recent enactment of a 14-day quarantine period for individuals entering Hong Kong from the mainland China, and the emergence of clustered local cases [19], the next important strategy on the agenda is to stabilize the supply of preventive materials, such as masks, so that the blockage of local transmission chain can be sustained. This is particularly important during the 24-day incubation period [20] associated with an elevated influx of individuals from the Mainland to Hong Kong between the announcement date (5 February 2020) and the effective date (8 February 2020) of the 14-day quarantine policy, and the recent emergence of super-spreaders to speed up local transmission.

Second, our results reveal the risk perception in the community, which is an important piece of information to enhance epidemic control [21]. Although the epicenter of the COVID-19 epidemic is Wuhan, the perceived risk of the community in Hong Kong was high. For emotional status, the HADS-A score in our survey (9.01) suggests that the community was borderline abnormal in terms of anxiety. Despite the slight difference in the inclusion of measurement items, the community was seemingly more anxious about the current COVID-19 epidemic (mean STAI score=2.00) than the 2009 pandemic influenza (mean STAI score = 1.8) [22], but was less worried than the SARS outbreak in 2003 (mean STAI score = 2.24) [23]. The significant time trend associated with HADS-A (Figure 2) suggests that the community became more and more anxious as new cases and new incidences came up (Figure 1).

Third, our results suggest an alternative strategy for better risk communication. The large proportion of respondents were alert to COVID-19 (99.5%) or actively searching for related information (83%) highlighted the role of social media in shaping risk perception and epidemic-related emotion. It is particularly important amid of much disease uncertainty as mass scares can be triggered easily. Considering the high level of trust given by respondents to doctors and the low level of trust to the two most frequently used information sources, social platform and websites, health officials can collaborate frequently with associations of medical doctors, and invite them to help propagating official information in more sociable channels. This strategy is deemed more acceptable by the community than relying solely on the official channel, given only 16% of respondents rates official websites as reliable or very reliable. Our results also shortlisted information preferred by the community among an upsurge of disease-related information during the early stage (Table 5).

Fourth, our results pinpoint the drivers for greater level of adoption of social-distancing precautionary measures. In line with literatures that being female and an elevated anxiety level prompted compliance of precautionary measures [23, 24], we also identified similar association in this survey (Table 6). Interestingly, specific to this COVID-19 epidemic, residents in the NT were more likely to comply with social-distancing precautionary measures than their counterparts in other areas of Hong Kong.

Separating Hong Kong and the Guangdong Province are two busiest custom borders, Lo Wu and Lok Ma Chau [19], such that the residents in NT may consider themselves at greater risk of infection. Those who claimed they understood COVID-19 were more likely to adopt preventive measures, suggesting mass promotion of knowledge about COVID-19 in the community can boost uptake of precautionary measures. On the other hand, the less propensity to adopt precautionary measures among individuals who left Hong Kong in the previous month or who regularly visited China reinforces the need for border screening and for promoting social hygiene amid of epidemic times.

Fifth, this local study has profound implication to overseas countries undergoing the initial phase of the COVID-19 epidemic. The WHO European region has been accumulating COVID-19 cases, but in only two days (22-24 February 2020), the number of laboratory-confirmed cases in Italy has risen from 17 to 219 [25]. Recently on 24 February 2020, the Ministry of Health announced the first COVID-19 case in Iraq. The presence of initial cases, aligning with the human-to-human [26] and asymptomatic [27] transmission, suggest that many countries may experience the initial phase of the COVID-19 epidemic soon. Results of this survey serve as a reference for overseas health officials to better prepare their containment strategies and handle the potential mass scares in their community.

This study has two strengths. First, it started within 36 hours after the detection of first local cases. This early start enables timely assessment of the community responses such that there is sufficient gap period to inform intervention policies. Second, our recruitment method, online survey via dissemination by DCCA councilors, is the first of its kinds to capture responses during public holidays while maintaining good geographical representation. The COVID-19 epidemic was amid of the Chinese New Year holidays and a series of large-scale social-distancing interventions enacted by Hong Kong government, particularly the home-office arrangement for employees. Therefore, the conventional random digit dialing approach adopted in the past local outbreaks [22, 23, 28] was not possible. And the involvement of all 452 DCCA councilors allows a thorough representation of every district in Hong Kong in the absence of a universal email database.

This study has two limitations. First, with an online approach, responses of those without internet access, particularly the oldest age group (55 years or above), were under-represented. Despite this, online surveys were the only feasible means of data collection during outbreak times. Second, this survey was conducted during the early phase that temporal variations of responses are not captured as the epidemic progresses. However, contact information were collected from this study cohort and follow-up surveys will be carried out as the disease progresses.

To conclude, we identified high risk-perception towards COVID-19 in the community, with the anxiety level higher than pandemic influenza but lower than SARS. Most respondents are alert to the disease progression of COVID-19, and adopt self-protective measures. This study contributes by examining the psycho-behavioral responses of hosts, in addition to the largely studied biological and mechanistic aspects of COVID-19, during the early phase of the current COVID-19 epidemic. The timely psychological and behavioral assessment of the community is useful to inform subsequent intervention and risk communication strategies as the epidemic progresses.

## Data Availability

The data will be available in github when it is officially published

